# A Web-based Tool for Automatically linking Clinical Trials to their Publications

**DOI:** 10.1101/2021.06.24.21259481

**Authors:** Neil R. Smalheiser, Arthur W. Holt

**Affiliations:** Department of Psychiatry, University of Illinois College of Medicine, 1601 W. Taylor Street, MC912, Chicago, IL 60612 USA

**Keywords:** clinical trials as topic, clinicaltrials.gov, evidence based medicine, bibliometrics, systematic reviews

## Abstract

**Objective:** Evidence synthesis teams, physicians, policy makers, and patients and their families all have an interest in following the outcomes of clinical trials and would benefit from being able to evaluate both the results posted in trial registries and in the publications that arise from them. Manual searching for publications arising from a given trial is a laborious and uncertain process. We sought to create a statistical model to automatically identify PubMed articles likely to report clinical outcome results from each registered trial in ClinicalTrials.gov.

**Materials and Methods:** A machine learning-based model was trained on pairs (publications linked to specific registered trials). Multiple features were constructed based on the degree of matching between the PubMed article metadata and specific fields of the trial registry, as well as matching with the set of publications already known to be linked to that trial.

**Results:** Evaluation of the model using NCT-linked articles as gold standard showed that they tend to be top ranked (median best rank = 1.0), and 91% of them are ranked in the top ten.

**Discussion:** Based on this model, we have created a free, public web based tool at http://arrowsmith.psych.uic.edu/cgi-bin/arrowsmith_uic/TrialPubLinking/trial_pub_link_start.cgithat, given any registered trial in ClinicalTrials.gov, presents a ranked list of the PubMed articles in order of estimated probability that they report clinical outcome data from that trial. The tool should greatly facilitate studies of trial outcome results and their relation to the original trial designs.

## OBJECTIVE

Clinical trials are the engine that drives improvements in health care. Evidence-based medicine seeks to collect and evaluate all possible evidence on a given question, giving highest priority to randomized controlled trials when available.[1] Evidence may reside in peer-reviewed publications that report trial clinical outcome data; clinical outcome data that is deposited in trial registries; grey literature; and patient-level trial data. Posted trial results often give more information about adverse events than those in the corresponding publications, and even such basic information as primary clinical outcome measures may differ between these sources.[2-6] Although it is unclear how often the conclusions of a systematic review are altered by including posted trial results,[7-9] physicians, policy makers, and patients and their families all have an interest in following the outcomes of registered trials,[10] and would benefit from being able to evaluate both the results posted in the trial registry and the publications that arise from them.

Finding the publications that arise from a given clinical trial is no easy matter, however, since only about half of trials give rise to any publications,[11-13] and of those that do, fewer than half mention the trial registry number to permit unambiguous linkage back to the trial.[14, 15] Manual searching for publications arising from a given trial is a laborious and uncertain process. [14, 15] Several machine learning methods employ textual similarity and other features to link publications to individual trials,[16-18] although there are currently no web based systems available for the biomedical community. In the present paper, we have created a free, public web based tool that, given any registered trial in ClinicalTrials.gov, presents a ranked list of the PubMed articles in order of the estimated probability that they report clinical outcome data from that trial.

## BACKGROUND AND SIGNIFICANCE

Linking publications to a given clinical trial is not a straightforward problem, for several reasons. First, the number of trials is large (∼390,000 in ClinicalTrials.gov as of December 1, 2020) and the number of potentially linked publications is even larger (∼32 million articles indexed in PubMed, of which ∼1.9 million articles mention the words trial or trials in title or abstract). Second, there is great variability among trials in the number of publications and their publication lags:[11, 12, 19-22] Although half of trials lack any publications, and most of those that do have only 1 or 2 publications, yet there is a long tail with some having >20 publications. Most are published between 2-5 years after the completion of the trial, but a few may be published after 10 or more years. Third, previously proposed methods of linking trials to publications (e.g., overall matching of textual similarity [18] or shared authors between trials and publications [14]) have limited predictive performance on their own. Fourth, the textual fields and metadata of trial registries are not well standardized,[17, 23, 24] which complicates the process of matching specific textual fields of trials to those of publications. Finally, ancillary publications may arise from a trial concerning a wide variety of issues, such as questionnaire development, GWAS studies carried out on trial subjects, reanalysis of data across multiple trials, and so on, which may not share word usage, topics, or investigators with the registered trial entry. Thus, similarity-based methods may be expected to be more successful for clinical outcome articles than for ancillary articles.

We built our Trials to Publications tool specifically for ClinicalTrials.gov and PubMed because they allow comprehensive and regularly updated XML-formatted downloading of all their trials and publications, which are not available for other trial registries and bibliographic databases. We have employed some of the features used in previous studies [17, 18] but have carried out additional trial-publication feature engineering, and have added a new feature based on the Aggregator model,[25, 26] which scores the degree of matching between a given candidate publication and the set of publications that are definitely known to be linked to the trial. As we will show, this substantially improves the performance of the model and surpasses previous efforts.

## MATERIALS AND METHODS

This section provides only a brief summary of methods. See the Supplementary File for full details regarding Methods.

### definitions and overview

Each registered trial in ClinicalTrials.gov is assigned a unique NCT number and has a trial registry entry consisting of multiple templated fields (e.g., start date, sponsor, inclusion and exclusion criteria, etc.). Publications that arise from a given trial are said to be “linked” to that trial, and comprise three different cases:

1. Some publications mention the NCT number explicitly in the abstract and/or are indexed in the PubMed record. Most of these are automatically recognized and posted as a templated field in the ClinicalTrials.gov registry. However, articles that only mention the NCT number within the full-text will be missed. As well, we have found that the automatic recognition system misses some textual variants of how the NCT number is written, as well as NCT numbers mentioned in the Corporate Author field; therefore, we have supplemented the automatically recognized set of NCT-linked articles with additional articles found using our own algorithms (Supplementary Methods). These were utilized as gold-standard training data for our modeling, excluding publications that contained multiple NCT numbers.
2. Some trials contain publications that were submitted by the trial investigators, but that do not contain NCT numbers. These comprise a heterogeneous set of articles (Figure 1); some provide clinical outcome results of the trial, but some are earlier studies or reviews which provided motivation for carrying out the trial. We attempted to identify the clinical outcome articles by making restrictions on publication date and requiring the article be listed in the specific results_reference field of clinicaltrials.gov (see Supplementary Methods for details). These were utilized as silver-standard training data for our modeling.
3. Finally, the goal of our modeling is to identify the set of PubMed-indexed publications that arise from a given trial, but that are not listed or explicitly attached to the ClinicalTrials.gov registry. The potential scope of our interest includes all PubMed articles -- whether or not they are clinical trial articles, and whether or not they have been assigned Medical Subject Headings or Publication Types.

**Figure 1.**
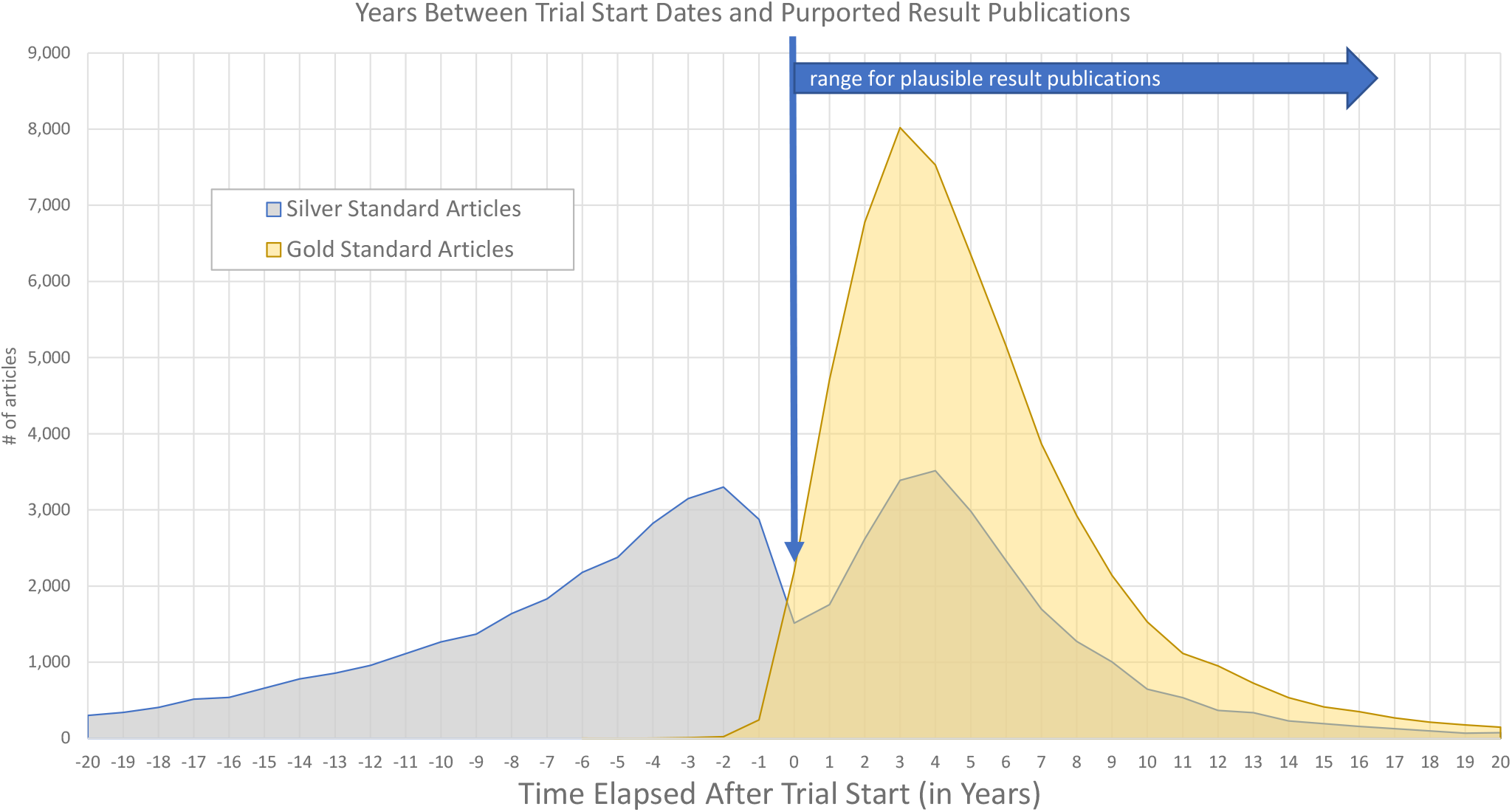
Relationship between trial start date and linked article publication dates for all registered trials in ClinicalTrials.gov. Data were collected in February, 2020. Publications with an explicit NCT link (shown in gold) most commonly appear 3 to 4 years after the start date of a trial. In contrast, investigator-submitted publications (shown in grey) exhibit a bimodal distribution of publication dates: Some are reviews or other publications that predate the start of a trial and provide motivation for carrying out the trial; in contrast, those published after the start of a trial appear to comprise primarily articles that arose from the trial itself.

### data preparation

We downloaded all public data as provided by the U.S. National Library of Medicine on the ClinicalTrials.gov website (https://clinicaltrials.gov/ct2/resources/download on 2/20/2020). The full dataset contained approximately 330,000 unique clinical trial registrations with all public data supplied in XML format. All xml fields were imported into a relational database for efficient data retrieval with a filter that retains only alpha-numeric characters and common punctuation. We also obtained PubMed records (metadata) readily available for all (then ∼30 million) articles at pubmed.gov.

### machine learning

Our strategy was to create positive training sets comprised of trial-publication pairs (using the gold-standard and/or silver standard data) vs. an equal sized negative training set constructed by randomly pairing trials with publications from other trials, matched such that they study the same condition or intervention. We extracted multiple features based on different aspects of matching between the trial and its linked publication, and used this to create a monotonic multi-dimensional measure of similarity that optimally distinguishes pairs in the positive vs. negative training sets. Finally, the similarity score between a trial and a publication is further modeled to estimate the probability that the publication arose from that trial (Supplementary Methods).

## RESULTS

The machine learning performance for the fitted model among the hold-out test cases, assuming a binary decision threshold of 0.5, was precision = 90.43% and recall = 84.57%, with F1 = 87.41%. Overall accuracy was 87.81% with an AUC of 0.95. These results, particularly the AUC value, indicate that the model is inherently able to discriminate positive vs. negative examples quite well.

However, it is more relevant to evaluate the model in the context of the implemented web-based ranking tool, which first identifies a pool of 5,000 candidate articles and then uses the model to make a ranked list according to their similarity scores. As gold standard, we employed NCT-linked articles but removed articles that were linked to more than one registered trial, as well as articles with publication dates prior to the trial start date. Note that this may provide an under-estimate of true performance, since it assumes that only the known NCT-linked articles are true positives; investigator-submitted articles, and true-positive articles that are not explicitly linked to them, are all counted as negatives in this evaluation. As discussed below, we carried out the evaluation separately for three cases: trials with two or more NCT-linked articles, trials with exactly one NCT-linked article, and trials with no NCT-linked articles.

### trials with two or more NCT-linked articles

In this situation, each test article is compared not only to the registered trial, but to each of the other articles known to be linked to the trial (including the NCT-linked articles as well as any investigator-submitted articles having publication dates later than the trial start date). A test article is not compared against itself, however. This situation provides the most accurate assessment of performance, including the contribution of the Aggregator feature, i.e., computing similarity between the test article and each of the articles known to be linked to the trial, which overall is the most powerful single feature in our model (Supplementary Methods).

As shown in Table 1, the model ranks NCT-linked articles extremely well, with a median first rank of 1.0 and median first similarity score of 0.993. Overall, about 78% of the NCT-linked articles arising from a trial are ranked in the top 10. (Note that the theoretical maximum recall is less than 100% for this category, since some trials have more than 10 linked articles.)

**Table 1.** Performance parameters for registered trials having ≥2 NCT-linked articles.

| Test Set | # Trials in Sample | Median First Rank (IQR) | MRR (95% C.I.) | Recall@5 (95% C.I.) | Recall@10 (95% C.I.) | Median First Score (95% C.I.) |
| --- | --- | --- | --- | --- | --- | --- |
| Trials with 2 or more NCT-linked articles | 300 | 1.0<br>(1.0-3.0) | 0.690<br>(0.618-0.765) | 0.660<br>(0.591-0.727) | 0.779<br>(0.719-0.841) | 0.993<br>(0.988-0.996) |
| Trials with 2 or more NCT-linked articles, without Aggregator feature | 300 | 2.0<br>(1.0-6.0) | 0.576<br>(0.498-0.657) | 0.534<br>(0.461-0.612) | 0.642<br>(0.566-0.717) | 0.951<br>(0.912-0.973) |

300 registered trials were randomly chosen that were known to be linked to 2 or more NCT-linked articles, and we applied the full model or the model lacking the Aggregator feature. The 5,000 candidate articles were all ranked but only the top 10 were displayed by the web-based tool. (See below for details.) The parameters include: Mean Reciprocal Rank, i.e., $1/(\text{mean rank across all linked articles})$ ; Recall@5, recall of the top 5 ranked articles; Recall@10, recall of the top 10 ranked articles; Median first rank, i.e. the top rank among all its NCT-linked articles; Median first similarity score, i.e., the top similarity score among all its NCT-linked articles; IQR, inter-quartile range of the median first rank scores across the dataset; 95% C.I., 95% Confidence Interval of the corresponding parameter. C.I. was calculated from a simple bootstrap of 1,000 samples with replacement.

If the Aggregator feature is removed entirely from the model, then the performance drops to a median first rank of 2.0 and median first score of 0.951 -- significantly lower (p = 1.04 × 10^−05^) but still quite respectable (Table 1).

### trials with exactly one NCT-linked article

Overall, trials in this situation (Table 2, row a) produce a ranked list of articles whose median first rank is 1.0 and median first similarity score is 0.993, essentially the same as observed above in the case of two or more NCT-linked articles. However, we were concerned that this might be an overestimate of the true performance since when there is only one article linked to a trial, the model allows that article to be compared with itself. In order to assess the performance a bit more realistically, we examined trials that had one NCT-linked article but also had one or more investigator-submitted articles -- in this situation, the test article is compared against the other investigator-submitted articles but <u>not</u> against itself. Table 2 row b shows that the median score of the NCT-linked article remains very high, 0.969.

**Table 2.** Performance parameters for registered trials having one NCT-linked article.

| Test Set | # Trials in Sample | Median First Rank (IQR) | MRR (95% C.I.) | Recall@5 (95% C.I.) | Recall@10 (95% C.I.) | Median First Score (95% C.I.) |
| --- | --- | --- | --- | --- | --- | --- |
| a) one NCT-linked article | 300 | 1.0 (1.0-1.0) | 0.851 (0.800-0.902) | 0.953 (0.910-0.990) | 0.983 (0.950-1.000) | 0.993 (0.988-0.996) |
| b) one NCT-linked article, but has other investigator submitted articles (only gold standard examples marked as positives) | 300 | 3.0 (1.0-14.0) | 0.492 (0.412-0.578) | 0.650 (0.560-0.740) | 0.730 (0.630-0.810) | 0.969 (0.951-0.982) |
| c) one NCT-linked article, but has other investigator submitted articles (gold and silver standard examples both marked as positives) | 300 | 1.5 (1.0-5.0) | 0.615 (0.537-0.693) | 0.533 (0.454-0.606) | 0.637 (0.561-0.707) | 0.984 (0.973-0.991) |
| d) one NCT-linked article, without Aggregator feature | 300 | 2.0 (1.0-23.0) | 0.506 (0.422-0.593) | 0.607 (0.520-0.700) | 0.690 (0.600-0.780) | 0.887 (0.753-0.942) |

Although the median rank of the NCT-linked article falls from 1.0 to 3.0, this effect is more apparent than real, since the investigator-submitted articles are competing for the top ranks, yet are not counted as positives in this Gold Standard evaluation. Table 2 row c corrects for this by counting both NCT-linked and investigator-submitted articles as positives, and in this case, the median first rank is 1.5 and median first similarity score is 0.984 -- this may be the fairest estimate of the performance of the model applied to trials that have a single NCT-linked article. As shown in Table 2 row d, if the Aggregator feature is removed entirely from the model, the performance falls substantially and with statistical significance (note the non-overlapping confidence intervals), with the median score of the NCT-linked article centered at 0.887 (compare with 0.993 observed in Table 2 row a).

### trials with no NCT-linked articles

This category comprises three different subcases: a) Roughly half of trials do not generate any publications at all, so for these the model could never hope to identify relevant outcome articles. b) An unknown percentage of trials generate publications that are not indexed in PubMed. Again, the model would not be able to find such articles. c) Finally, of greatest interest, a minority of trials generate PubMed articles that are not identifiable by ClinicalTrials.gov nor our own scraping efforts, because they do not specify NCT numbers in the abstract or record metadata. Some of these can be verified by manual inspection of full-text (i.e., the NCT number is sometimes given in the Methods section), whereas others are not clearly linked to a specific registered trial even after inspecting the full-text.

We created a random sample of 100 registered trials lacking any NCT-linked and investigator submitted articles, made a ranked list of 5,000 PubMed articles for each trial, and plotted the best predictive score for each trial. As shown in Figure 2, only 12 of the 100 trials had best similarity scores above 0.98, in contrast to 79 of 100 randomly chosen trials that had two or more NCT-linked articles (Figure 2). Examining the best-scoring article in the 9 trials having scores above 0.99, two were definitely linked to the trial in question (the NCT number was listed within the full-text) and two probably belonged (same topic, investigator and institution). Two were associated with different trials (different NCT number given in full text) but were closely related, for example, the same investigator studying “”Hepatic Function During and Following Three Days of Acetaminophen Dosing vs. “Aminotransferase Trends During Prolonged Acetaminophen Dosing”. This suggests that screening trials with no known linked articles for very high-scoring candidate articles can find at least some true positives or those associated with closely related trials.

**Figure 2.**
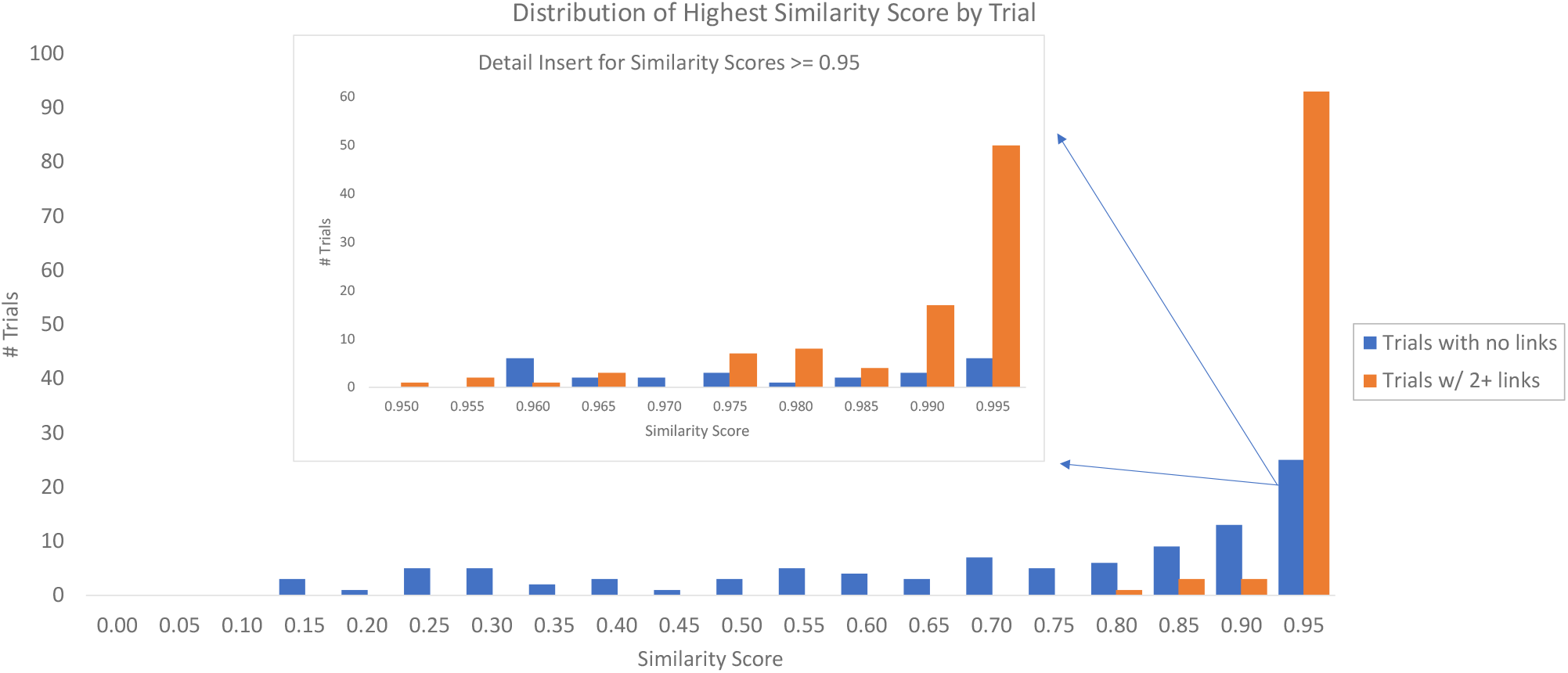
Best model-derived similarity scores for ranked article lists processed for 100 trials with no known results publications vs. 100 trials with 2 or more NCT-linked publications.

### comparison of our model to previous published automated methods

Goodwin et al has published a deep learning-based, multi-feature model to identify publications that are linked to registered trials.[17] It is not possible to make a direct comparison between our model and theirs, because their system is not currently available, and because their reported performance parameters are not based on the same corpus of trials or articles as ours. Nevertheless, we computed the same information retrieval performance metrics as Goodwin et al did, in order to make an approximate comparison of methods. Table 3 row b shows the performance of our most accurate evaluation situation (i.e., trials with two or more NCT-linked articles), compared to Goodwin’s best and most comparable evaluation situation (Table 3 row a). Our method exceeds substantially all parameters compared to Goodwin et al.

**Table 3.** Performance parameters to compare our methods vs. Goodwin et al [17].

| Reported Model Version | MAP | MRR | R-Prec | P @ 5 | P @ 10 |
| --- | --- | --- | --- | --- | --- |
| a) Goodwin et al., Closed Strategy<br>NCT Link DHN | 0.308 | 0.342 | 0.244 | 0.123 | 0.082 |
| b) Trials with 2 or more NCT-linked<br>articles | 0.591<br>(0.524-0.659) | 0.690<br>(0.618-0.765) | 0.506<br>(0.431-0.582) | 0.342<br>(0.304-0.380) | 0.213<br>(0.191-0.238) |

Shown are our performance parameters compared with those reported by Goodwin et al for their best and most comparable method.[17] MAP, Mean Average Precision; R-Prec, R-Precision; P@5, P@10, Precision of the top 5 and 10 ranked articles.

Dunn et al have also published a trials to publications model based on textual and conceptual similarity between article and trial metadata.[18] Their code is archived publicly at https://github.com/pmartin23/tfidf and so we were able to compare their methods directly with ours. We used the same test corpus of 300 trials as reported in Table 1 (except that a few articles could not be parsed by Dunn’s system and were removed from our evaluation as well). As shown in Table 4, our performance was greater for all parameters, and the difference was statistically highly significant. Further analysis (see above and data not shown) suggests that our improved performance is largely due to the Aggregator feature, which complements textual and semantic similarity measures.

**Table 4.**
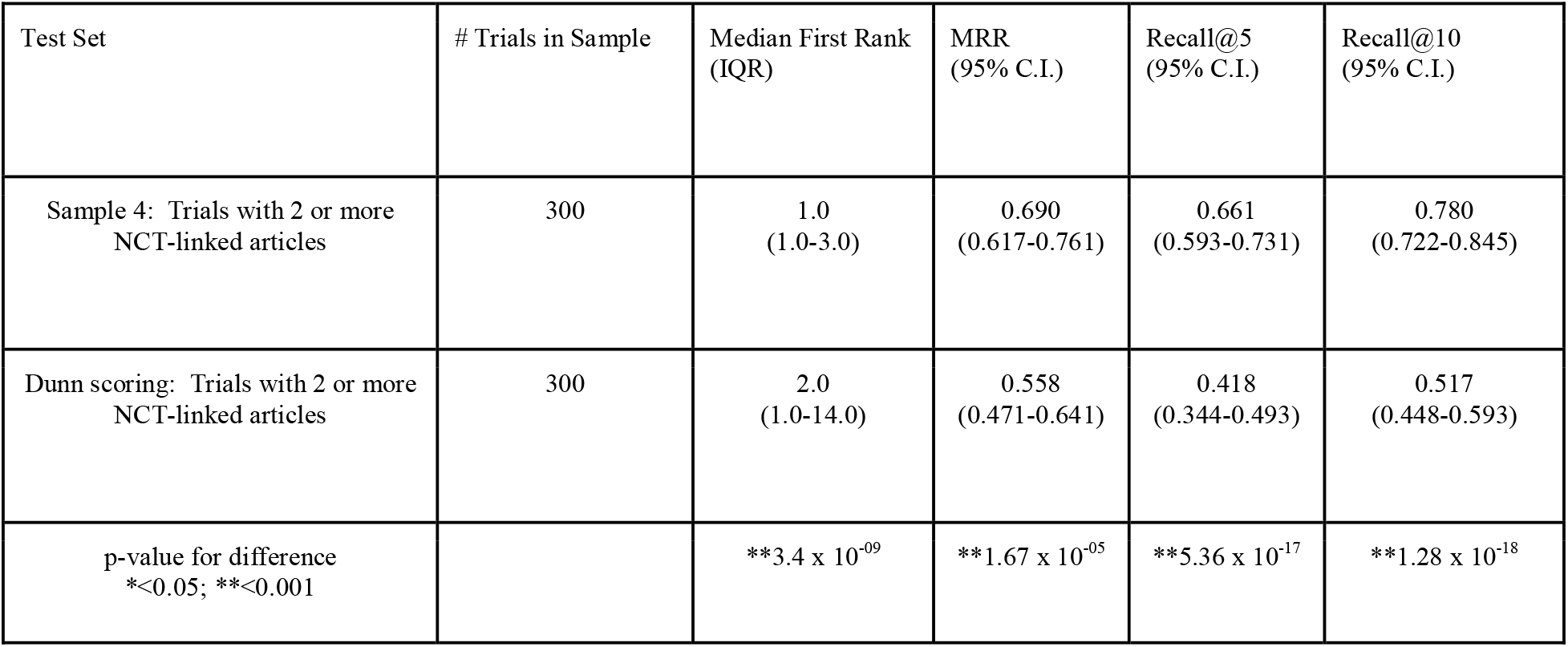
Performance parameters to compare our methods vs. Dunn et al [18].

| Test Set | # Trials in Sample | Median First Rank (IQR) | MRR (95% C.I.) | Recall@5 (95% C.I.) | Recall@10 (95% C.I.) |
| --- | --- | --- | --- | --- | --- |
| Sample 4: Trials with 2 or more NCT-linked articles | 300 | 1.0<br>(1.0-3.0) | 0.690<br>(0.617-0.761) | 0.661<br>(0.593-0.731) | 0.780<br>(0.722-0.845) |
| Dunn scoring: Trials with 2 or more NCT-linked articles | 300 | 2.0<br>(1.0-14.0) | 0.558<br>(0.471-0.641) | 0.418<br>(0.344-0.493) | 0.517<br>(0.448-0.593) |
| p-value for difference<br>*$<0.05$; **$<0.001$ | | **$3.4 \times 10^{-09}$ | **$1.67 \times 10^{-05}$ | **$5.36 \times 10^{-17}$ | **$1.28 \times 10^{-18}$ |

### error analysis

We manually examined a selection of articles that were given predictive scores >0.99 yet definitely did not arise from the registered trial as predicted (e.g., associated with different NCT numbers). The most common source of errors were caused by investigators who have individually produced numerous trials and numerous publications regarding the same condition or treatment. Thus, articles may be predicted to belong to one registered trial when, in fact, they belong to a related or follow-up trial (e.g., a phase III trial rather than phase II). A similar type of error could also occur for heavily studied conditions (e.g., metformin to treat diabetes) in which many similar trials produced many similar publications. Even these errors do not entirely reduce the utility of our tool, however: Since most trials generate only a few publications at most, displaying the top 10 publications (along with any that explicitly list NCT or other trial registry numbers) should point to the most related trials.

### implementing the tool based on the model

A web-based query interface that implements our model is shown in Figure 3. The user is prompted to enter a valid NCT number of a registered trial in ClinicalTrials.gov. In the basic search mode, the model is applied to a preselected list of 5,000 candidate PubMed articles based on shared conditions or interventions and/or some degree of textual similarity (Supplementary Methods). The advanced search interface allows the user to specify which PubMed articles should be processed; one can retrieve up to 100,000 articles that satisfy a user-specified PubMed query (Supplementary Methods). In the back end of the web service, our database is automatically incremented for newly registered trials and newly published articles on a weekly basis.

**Figure 3.**
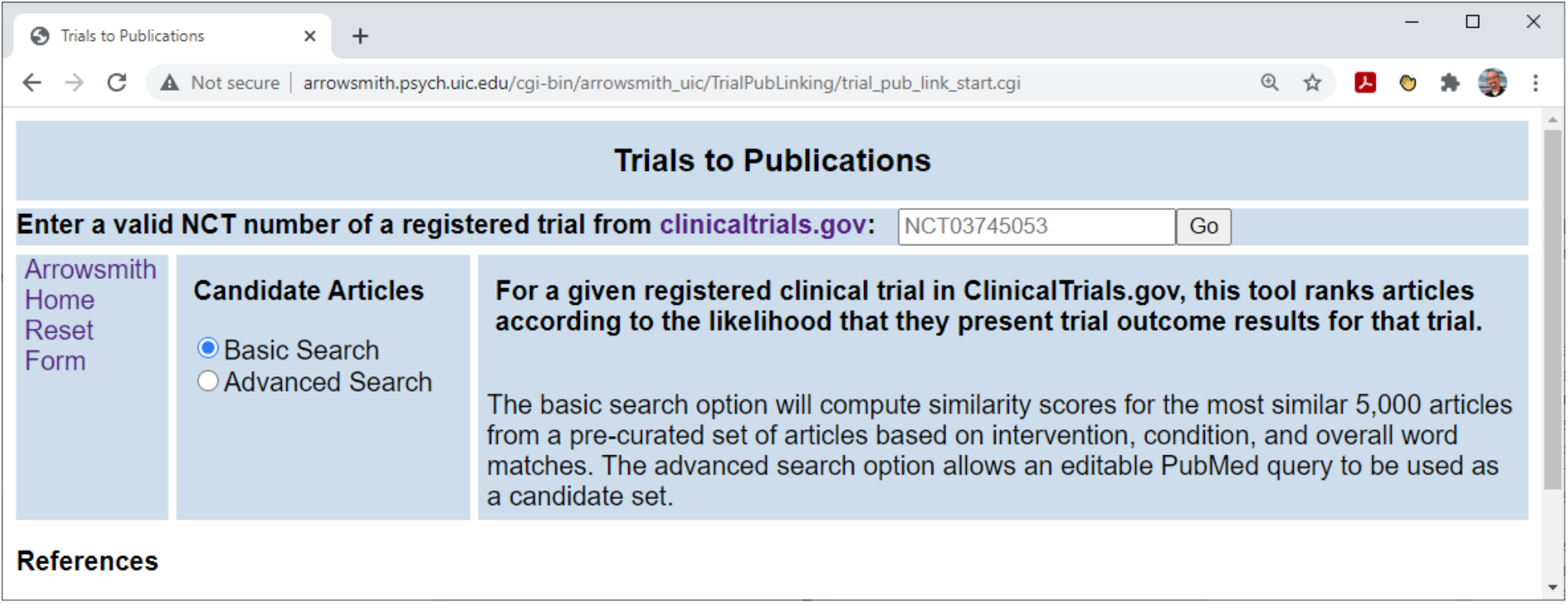
Screenshot of the Trials to Publications query interface. Visitors may view top-ranked articles for any valid ClinicalTrials.gov NCT number with a predetermined candidate set (Basic Search) or a PubMed.gov compatible query (Advanced Search). In the Advanced Search, the user is offered a PubMed query that is pre-populated with suggested terms taken from the condition, intervention and investigator fields of the registered trial, but can be freely edited so that, in effect, the user can enter any PubMed query at all, and create a candidate set of PubMed articles of possibly any size. This allows maximal flexibility. However, some guidance will be required since such queries cannot be pre-calculated but must be run in real time, and large sets may potentially take hours to process.

As shown in Figure 3, the user enters a valid NCT number of a trial registered in ClinicalTrials.gov, and receives a list of PubMed articles ranked according to similarity score (Figure 4). Note that in the Basic Search, we have restricted the pool of candidate articles to those which were published after the trial start date, and that share at least some minimal features with the registered trial (Supplementary Methods). If more than 5,000 articles satisfy these criteria, then the pool is limited to the 5,000 which match best on the initial feature set. A pool of 5,000 articles can be scored and ranked in real time within ∼10 minutes. Because most trials have no or very few publications associated with them, displaying the top 10 articles will capture nearly all relevant articles in most cases. However, if all 10 have similarity scores >0.8, then the display is extended to show all articles that have scores ≥ 0.8. In addition to displaying the similarity score for each article, we also display the estimated probability that the article arose from the given trial (see Figure 4 and Supplementary Methods). Note that probabilities are not simply proportional to similarity scores, so that a similarity score of 76.3% implies only a 1.9% chance that the article arose from that trial (Figure 4). Articles that share the same NCT number as the trial are displayed at the top of the page, along with investigator-submitted articles, regardless of their similarity score. Finally, we have scraped registry numbers for a large number of international trial registries (Supplementary Methods); if a ranked article is linked to any of these registries, we display the registry number next to the article.

**Figure 4.**
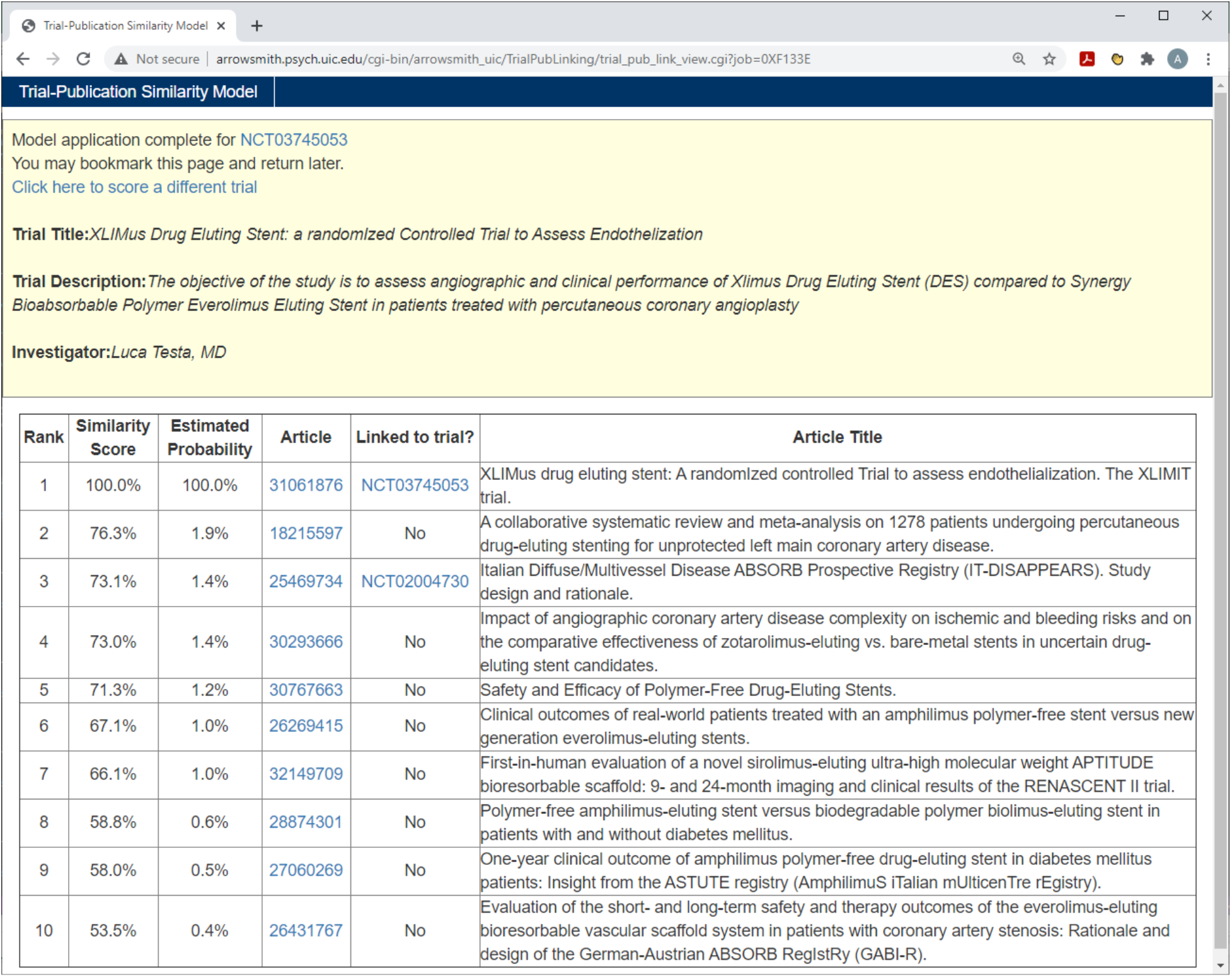
Screenshot of the Results page for the query shown in Figure 3.

## DISCUSSION

We present a machine learning based model and web-based tool that automatically predicts, for any given registered trial in ClinicalTrials.gov, which PubMed articles are the most likely to present its clinical outcome results. The underlying assumption is that articles that describe clinical outcomes from a trial will tend to share similarities with the trial registry in terms of text, MeSH terms, and/or investigator names. In addition, we introduced an additional type of “Aggregator” feature [25, 26], in which articles were also compared for similarity with any articles known to be linked to the trial by virtue of having listed explicit NCT numbers or having been manually submitted by the trial investigators. The similarity metrics were trained using a corpus of positive and negative examples taken from ClinicalTrials.gov, and evaluated on different test sets of articles and trials.

Our evaluations, using NCT-linked articles and investigator-submitted articles as gold and silver standards, verified the similarity assumption and showed that the model placed the majority of known linked articles in the top 5. Conversely, articles that had extremely high similarity scores with a given registered trial (e.g., >0.99) often arose from that trial. Direct comparisons with previous efforts suggest that our method exceeds the current state of the art, and that the “Aggregator” feature contributes significantly to the overall performance. The web-based tool is free and publicly available at http://arrowsmith.psych.uic.edu/cgi-bin/arrowsmith_uic/TrialPubLinking/trial_pub_link_start.cgi.

Several important limitations should be noted. Our tool is restricted to matching ClinicalTrials.gov with PubMed articles, so it does not include other registries or other bibliographic databases. Although the matching is based on multiple aspects of similarity, which should capture most articles that present clinical outcome results, we will miss a small proportion of articles that arise from the trial, such as questionnaire development studies. Preselecting 5,000 candidate articles reduces the time for presenting ranked results to under 10 minutes at present. However, we are currently in the process of pre-processing all existing trials and their candidate publications, which will allow almost instantaneous display in most cases.

## CONCLUSION

The ability to find articles that are closely related to a given registered trial should provide value for physicians, patients and their families who seek to follow up on individual trials, as well as evidence synthesis teams seeking to find relevant publications. Future refinements to the tool, for example the Advanced search option, will depend on user feedback. Depending on the interest from the research community, we may seek to learn if there is sufficient interest to engineer the tool in the reverse direction, that is, given a PubMed clinical trial article, to identify its most similar registered trials.

## Data Availability

The web-based tool is free and publicly available at http://arrowsmith.psych.uic.edu/cgi-bin/arrowsmith_uic/TrialPubLinking/trial_pub_link_start.cgi.

## Acknowledgements

Thanks to Adam Dunn and Shifeng Liu for their cooperation and advice in evaluating their previously published model in comparison with ours.

## Supplementary File

This file contains the details of methods employed in our research and implementation of the web-based tool.

## AUTHOR CONTRIBUTIONS

Neil Smalheiser: conceptualization, methodology, writing-original draft, writing -review and editing, supervision, funding acquisition.

Arthur Holt: methodology, software, validation, formal analysis, investigation, writing - original draft, writing -review and editing, visualization.

