## Supplementary File for "A Web-based Tool for Automatically linking Clinical Trials to their Publications"

1. **SUPPLEMENTARY METHODS**

**1.1 Creating a Gold Standard set of trial-article pairs**

Although ClinicalTrials.gov has a procedure for automatically detecting mentions of registry (NCT) numbers within PubMed article metadata, we found that some articles were missed due to textual variations or placement of the NCT number into fields such as Corporate Author. Thus, we developed our own search rules to create a dataset of NCT registry numbers mentioned within an article’s Title, Abstract, Accession Number and Corporate Author fields of PubMed article XML metadata.  For all PubMed articles written in English, we searched for the stem “NCT”, followed by a series of numbers, while tolerating mixed capitalization and the occurrence of one extra space or other character between “NCT” and the numeric digits using the python-flavor regular expression  '(NCT.?[0-9]+)'.  As of January 2020, we found 57,575 Pub-Med articles that contained an explicit and valid NCT number as determined by our text scraping process, and denote these trial-article pairs as our gold standard positive cases.  In addition to and distinct from these links, we identified 61,099 trial-article pairs that were listed in the “results reference PMID” XML metadata from ClinicalTrials.gov, which we designate silver standard positive cases.  These articles were submitted by the trial investigators; some of them present results from the trial, whereas some articles predate the trial and are meant to provide motivation for conducting the trial. As explained in MATERIALS AND METHODS, we filtered these articles by publication date to create a less noisy set of clinical outcome results publications.

**1.2 Defining and repairing start dates and publication dates for trials and articles**

ClinicalTrials.gov has multiple fields for defining start dates -- first posted date, first submitted date, trial start date and verification date -- that do not necessarily agree closely with each other. Similarly, PubMed articles have several fields for publication date (e.g., first posted online vs. in print), and dates may be given precisely (to a specific date) or only to a season (e.g., Summer). Thus, we designed methods to repair article publication dates, trial start dates, and trial completion dates to enable date differential computations and comparisons.  To normalize publication dates as they are represented in the Publication Date (DP) PubMed field, we corrected dates containing only a season by substituting the middle month of the season (e.g. substituting July for “summer”), and substituting June for articles having only a publication year.  We assigned the 15th of the month when necessary to form a complete date.

For registered trials in ClinicalTrials.gov, the trial completion date and trial start dates most commonly failed conversion to a numeric date, i.e. “rogue values”, when the day of the month was missing.  We repaired these dates by inserting the 15th of the month, as with article publication dates.  After examination of the differential, in years, between NCT-linked article publish dates and both trial start date and trial completion date, we determined that the trial start dates provide a more reliable time-based filter to reduce the population of possibly relevant articles that may contain trial results (Figure 1 in main text). A detailed review of  34,570 known trial-article links, restricted to randomized controlled trial articles, uncovered only 206 trial-article pairs for which our repaired publication date precedes our repaired trial start date.  Of these, 173 fell within -1 years and mostly resulted from our substitution of June 15th when our publication month was missing.  All but 13 could be attributed to long delays between a trial’s posting on clinicaltrials.gov and the trial start date.  Each of these 13 “errors” were determined to be irreparable date formats or incorrect NCT numbers found in the article abstracts.

**1.3 Development of Model Sample**

We constructed a modeling dataset with binary positive and negative labels for classification in a manner that was designed to generate negative cases of trial-article pairs that are at least somewhat related to each other.  Our list of positive cases began with all trial-article pairs for which explicit NCT number tags are found in the article meta-data (our gold standard).  We further restricted positive cases to articles that contained the words “trial” or “trials” or “multicenter study” and were published at least one year after the trial start date, or 0 years after the trial start date if the article publishing date had only been represented as a year.  Also, we removed articles that mentioned more than one trial NCT number, and allowed no more than 3 articles from any one trial, so that our dataset would not be unduly biased towards trials that generated many publications.  There remained 21,430 positive trial-article pairs in the NCT-linked (gold standard) training set.

Negative cases were constructed with a custom algorithm that selects trial-article pairs which may be at least partially related to each other.  This was done to be consistent with how we anticipate our model would be applied in the real-world, in which searches will generally be constrained to studies of a particular condition or intervention.  We selected potential negative trial-article pairs as follows:

- For each trial in our positive set, choose all other trials with their NCT-linked articles such that:
  - All Medical Subject Headings (MeSH terms) from the Condition field of one trial appear in those of the other trial
  - OR all trial intervention MeSH terms from one trial appear in those of the other trial
  - “Placebo” and “Healthy” MeSH terms were ignored for this purpose.
  - No more than 3 trial-article pairs in the negative set may come from any one trial

.

This generated over 7 million potential negative pairs, from which we randomly sampled 21,430 to provide equally sized positive and negative training pairs.

Positive/Negative Pairs

Trial

Publication

Explicit Link

(NCT number in article meta-data)

Trial

Publications

Conditions

OR

Interventions

Positive

Negative

Trials

**Figure S1: positive and negative case derivation.** Positive cases consist of “clean” NCT-linked articles allowing no more than 3 from a single trial.  Negative article cases are designed to be at least somewhat related to the paired trial.

**1.4 Feature engineering**

We selected templated fields from trials and articles for matching, taking inspiration from Goodwin et al [1], while adding additional comparisons hypothesized or empirically suggested to be important by simple shared word count indexing among known trial-article links.  For each potential comparison between trial text and article meta-data, we applied one or several comparative methods.  For comparisons of English-like text, we computed weighted and unweighted shared terms and implicit scores as described in [2], as well as Okapi BM25 similarity scores.  For comparisons involving authors and investigators, we applied the Author-ity author name matching algorithm [3] that assigns a score from 0 to 100 determined by levels of partial match for last name and initials, with human names parsed from the trial XML elements with the python Nameparser library [4].  For data elements that generally contained IDs or lists, such as MeSH terms, we computed simple occurrence counts and/or the proportion of items in the list found.  For MeSH terms in particular, we tested several versions of comparison including the proportion of article MeSH terms found and the arithmetic and geometric mean of the derived odds for all possible pairs of terms from the two lists as described in [5].  These variants showed no improved predictive utility over the sum of the proportions for both directions, that is:  (proportion of article MeSH terms listed in the Trial MeSH terms) + (proportion of Trial MeSH terms appearing in the article MeSH list).  Table S2 contains a list of all combinations of trial registry field attributes, PubMed article metadata fields, and matching algorithms that were considered as potential predictive features.

**Table S2:  Trial and Article Data Comparisons Considered in Constructing the Model.**

| Trial Attribute | Trial XML Tag | PUBMED Fields | Comparison Methods |
| --- | --- | --- | --- |
| Title | //clinical_study/brief_title | Title+Abstract | BM25\|Tsim\|Allcaps |
| Sponsor Name | //clinical_study/sponsors/lead_sponsor/agency | Affiliation\|CN | BM25\|occurrence |
| Official Title | //clinical_study/official_title | Title+Abstract | BM25\|Tsim\|Allcaps |
| Brief Summary | //clinical_study/brief_summary/textblock | Title+Abstract | BM25\|Tsim\|Allcaps |
| Study Description | //clinical_study/detailed_description/textblock | Title+Abstract | BM25\|Tsim\|Allcaps |
| Eligibility Criteria | //clinical_study/eligibility/study_pop/textblock | Title+Abstract | BM25\|Tsim |
| Inclusion | //clinical_study/eligibility/criteria/textblock | Title+Abstract | BM25\|Tsim |
| Exclusion | //clinical_study/eligibility/criteria/textblock | Title+Abstract | BM25\|Tsim |
| Agency Class | //clinical_study/sponsors/collaborator/agency_class | Grants\|CN\|Affiliation | BM25 |
| Investigator | //clinical_study/overall_official/last_name | Authors | Authtree |
| Investigator full name | //clinical_study/responsible_party/investigator_full_name | Authors | Authtree |
| Results Contact | //clinical_study/clinical_results/point_of_contact/name_or_title | Authors | Authtree |
| Study Type | //clinical_study/study_type | Title+Abstract | Occurrence |
| Allocation | //clinical_study/study_design_info/allocation | Title+Abstract | Occurrence |
| Primary Purpose | //clinical_study/study_design_info/primary_purpose | Title+Abstract | Occurrence |
| Conditions | //clinical_study/condition | Title+Abstract | propoccurrence |
| Interventions | //clinical_study/intervention/intervention_name | Title+Abstract | propoccurrence |
| Phase | //clinical_study/phase | Title+Abstract | Occurrence |
| Org Study ID | //clinical_study/id_info/org_study_id | Title\|Affiliation\|Abstract | Occurrence |
| Affiliation | //clinical_study/overall_official/affiliation | Affiliation\|CN | Occurrence |
| Country | //clinical_study/location_countries/country | Title\|Grants\|Abstract\|Affiliation | Occurrence |
| Acronym | //clinical_study/acronym' | Abstract\|CN | Occurrence |
| Secondary ID | //clinical_study/id_info/secondary_id | Grants\|Affiliation\|CN | Occurrence |
| Investigator | //clinical_study/overall_official/last_name | Author Email | Reverseoccur |
| Investigator full name | //clinical_study/responsible_party/investigator_full_name | Author Email | Reverseoccur |
| Primary Purpose | //clinical_study/study_design_info/primary_purpose | NM | Reverseoccur |
| Official Title | //clinical_study/official_title | NM | Reverseoccur |
| Brief Summary | //clinical_study/brief_summary/textblock | NM | Reverseoccur |
| Study Description | //clinical_study/detailed_description/textblock | NM | Reverseoccur |
| Interventions | //clinical_study/intervention/intervention_name | NM | Reverseoccur |
| Linked MeSH Terms | //clinical_study/intervention_browse/mesh_term | NM | Reverseoccur |
| Results Contact | //clinical_study/clinical_results/point_of_contact/email | Author Email | Reverseoccur |
| Linked MeSH Terms | //clinical_study/intervention_browse/mesh_term | MeSH | MeSHsim |
| Condition MeSH | //clinical_study/condition_browse/mesh_term | MeSH | MeSHsim |

**Complex text similarity metrics such as BM25 and word vector comparisons were calculated among free-form text fields**.  Item lists such as MeSH terms were compared with proportions of items appearing in the trial or article fields.  Codes and identifiers were represented with a simple indicator if they appeared in the article metadata.  Comparison methods are defined as follows:

Tsim - text similarity [4]. We examined a variety of text similarity measures, including Unweighted Implicit Shared Terms, Weighted Implicit Shared Terms, Implicit Weighted Score, and Implicit Unweighted Score.

BM25 - Okapi BM25 similarity.

Occurrence - simple count of unique trial words occurring in article field.

Reverseoccur - simple count of publication words occurring in trial field.

propoccurrence - proportion of terms in a list found in a comparison text.

MeSHsim - the proportion of MeSH terms matching after stop listing the 20 most frequent MeSH terms in MEDLINE, summed across both directions.

Authtree - the author name partial matching algorithm from the Aggregator model [3], with HumanName parsing.

Allcaps - the number of matching all-capitalized words with a minimum of 3 letters after stoplisting with the PubMed 365 word stop list.

**1.5 Trial and Article Dates**

Multiple differentials between trial dates and article publication dates were tested for efficacy as a feature to predict the likelihood of a trial-article match.  Aside from a few cases of rogue date formats [1,6], the primary and secondary trial completion dates were unreliable since trials may continue for decades and it is not unusual for the purported trial completion date to be in the distant future whilst linked articles may be published with interim results.  We found the discriminant ratio between the start date as provided by the //clinical_study/start_date XML field from ClinicalTrials.gov vs. the article PD publication date, both repaired as described above, to be the most stable and predictive feature for linear modeling.

**1.6 Aggregator Relational Features**

We introduce a novel class of features that provide excellent predictive utility when they may be computed.  Recall that many registered trials contain one or more known publications x1, x2, x3, … that are explicitly linked to that trial, either as determined by our gold standard NCT link or silver standard investigator-submitted results link  Thus, when one is considering a candidate pool of PubMed articles y1, y2, y3... to be compared and ranked for similarity to the registered trial, one can consider similarity features between the trial and each of the candidate PubMed articles yi. As well, one can consider similarity features between each candidate PubMed article yi and the set of known publications xi associated with that trial. For any PubMed article x1 that is being considered for its similarity to a given registered trial, we employed the Aggegator pairwise similarity model [7, 8] to calculate similarity metrics between that article and all *other* articles known to be associated with the same trial. We found the maximum of all the pairwise x1-yi similarity scores to be the best performing feature and superior to the arithmetic mean and median.

Several other variations of the related article ranking features from the Aggregator model were computed and evaluated as separate features.  For each trial-article pair, we collected the values of the related article rankings from PubMed from the perspective of the article being considered to each of the known NCT linked articles for the trial.  We retained the average and minimum of these rankings for testing as potential predictors.

These other-linked-article features provided a substantial contribution to our model, but also introduced several complications:

1) Aggregator scores are highly correlated with shared Investigators and Authors among articles.  To resolve the multicollinearity between Aggregator scores and Investigator-Author matches, our final model combines both features with a custom interaction term.  From our model training dataset, we developed a translation between the number of shared Investigator and Author names and the Aggregator score feature for cases where both features were available.  The combined feature takes on the maximum non-missing value as provided by either Investigator/Author matches or Aggregator.  If neither feature can be computed (i.e. no Investigator or Author data and no linked publications), a mean substitution is used.

2) Trials with no known linked articles do not benefit from the Aggregator feature.

3) Trials with only one known linked article are problematic for evaluating model performance, since the same NCT-linked article is used for modeling and for evaluation. Special considerations were taken to estimate performance in an unbiased manner (see Results in main text).

Missing values were handled in a manner that preserves the distinction between missing values that arise from a comparison not finding matching items between a trial and an article, versus missing values that arise from a trial or article data element not existing.  For features that count the occurrence of lists of items (e.g. article MeSH terms in a trial description), we recode the feature to be zero if any component of the feature did not exist, negative 0.5 if all components existed yet no match was found, and positive 0.5 if a match was found.  This treatment effectively capped the occurrence features to count only one match since, in real-world testing, we found that these features were unstable in that a few articles may happen to repeat a matching word many times in the title or abstract.  We combined all three of our article author and trial investigator features, namely article authors compared to each of trial Overall Official, Investigator Full Name and Results Contact, into a single feature taking the maximum of non-missing values.  This was done to avoid implementing an error prone process of deduplicating investigator names within a trial record.  When all three author derived features were missing as well as our Aggregator computation, we assign a mean substitution for the combined feature, as described above.  All remaining features received a simple mean substitution computed from our model training data.

**1.8 Feature selection and optimizing the fit of the model**

We chose to fit a classical Logistic Regression model because of its interpretability and ease of implementation, and because we expected no strong nonlinearities or nonseparable feature interactions. Thirty percent (30%) of our modeling dataset was held out for use as a test sample leaving us with 29,818 model training cases and 13,042 cases for model validation, both balanced with equal positive and negative trial-article pairs.  Model development began with a forward stepwise process with features added until the model adjusted r-square no longer increased.  We further optimized the model and reduced feature multicollinearity by choosing only the best performing comparison method for each trial-article pair of elements when multiple methods had entered the model via forward selection.  Other model adjustments were selected by careful examination of model performance and specific errors.  The covariance between our Aggregator feature and author/investigator comparison features was addressed with the custom interaction term described above.  Features that compared trial sponsors with article affiliations were predictive, but extremely error prone as described in [1, 6].  After extensive experimentation attempting to mute the effect of erroneous and mis-matched organizational names, we chose to exclude the affiliations features from the model.

**Table S5:  Regression Model Parameters (model with Logit link function)**

| Feature | Coefficient | Std Err | z | p>\|z\| | Lower 95% CI | Upper 95% CI |
| --- | --- | --- | --- | --- | --- | --- |
| Model Intercept | -3.516664 | 0.144 | -24.356 | 0 | -3.8 | -3.234 |
| Investigator/Aggregator Max | 7.064431 | 0.103 | 68.377 | 0 | 6.862 | 7.267 |
| Trial Interventions with article titleabs using propoccurrence | 1.321135 | 0.042 | 31.717 | 0 | 1.239 | 1.403 |
| Trial Brief Summary with article titleabs using tsimw_scr | 0.000237 | 0.000009 | 26.405 | 0 | 0 | 0 |
| Trial Intervention MeSH with article mesh using meshsim | 1.54084 | 0.064 | 24.079 | 0 | 1.415 | 1.666 |
| Discrim Ratio:start date | 1.03154 | 0.059 | 17.427 | 0 | 0.916 | 1.148 |
| Trial Acronym with article abstract using occurrence | 1.371825 | 0.083 | 16.56 | 0 | 1.209 | 1.534 |
| Trial Condition MeSH with article mesh using meshsim | 0.794547 | 0.05 | 16.025 | 0 | 0.697 | 0.892 |
| Trial Title with article titleabs using allcaps | 0.805654 | 0.052 | 15.402 | 0 | 0.703 | 0.908 |
| Trial Country with article grants using occurrence | 1.19076 | 0.08 | 14.968 | 0 | 1.035 | 1.347 |
| Trial Secondary ID with article grants using occurrence | 2.002089 | 0.137 | 14.581 | 0 | 1.733 | 2.271 |
| Trial Title/Desc and Article Title/Abs All Cap Mismatch | -1.290924 | 0.102 | -12.634 | 0 | -1.491 | -1.091 |
| Trial Conditions with article titleabs using propoccurrence | 0.50953 | 0.04 | 12.605 | 0 | 0.43 | 0.589 |
| Trial Org Study ID with article abstract using occurrence | 2.118923 | 0.177 | 11.964 | 0 | 1.772 | 2.466 |
| Trial Brief Summary with article chemicals using reverseoccur | 2.641784 | 0.228 | 11.596 | 0 | 2.195 | 3.088 |
| Trial Allocation with article titleabs using occurrence | 0.412927 | 0.043 | 9.703 | 0 | 0.33 | 0.496 |
| Trial Acronym with article cn using occurrence | 2.980237 | 0.336 | 8.871 | 0 | 2.322 | 3.639 |
| Trial Study Type with article titleabs using occurrence | 1.448419 | 0.165 | 8.765 | 0 | 1.125 | 1.772 |

The z-scores associated with a test for equality with zero for our final model parameters are shown in Figure S5.  As noted above, our Aggregator feature that compares prospective articles to those that are known to be linked to each trial is our strongest predictor (has the greatest coefficient).  After that, our top 5 predictors include two versions of trial intervention matching, the weighted text similarity score [2] between the trial summary and article title/abstract, and the time differential from trial start date.  The remaining 12 features serve primarily as indicator variables that provide support for article ranking for trials; these may be most useful in cases that do not have known article links.

**1.9 Converting the model’s raw similarity score to an estimated probability value, by using oversampled Odds Ratio smoothing**

Given that our model is derived from a balanced sample of positive and negative cases, we engineered a simple function to transform model scores to estimated probabilities.  We quantize the model predicted probabilities from our modeling dataset to the nearest hundredth to form 100 buckets from 0.00 to 0.99.  For the observed positive odds ratio, we smoothed the positivity rate across buckets with 3 simple linear segments from .00 to .75, from .76 to .96, and .97 to 1.00, and reverted them back to an odds ratio for each bucket (Figure S6). We then adjust the odds ratio for each bucket, effectively treating each model score bucket as an individual oversampled set, and convert to a probability for display in our web tool.

adjusted odds for bucket i = smoothed odds ratio for bucket i odds positive case appears in bucket i

adjusted probability for bucket i =11+1adjusted odds for bucket i

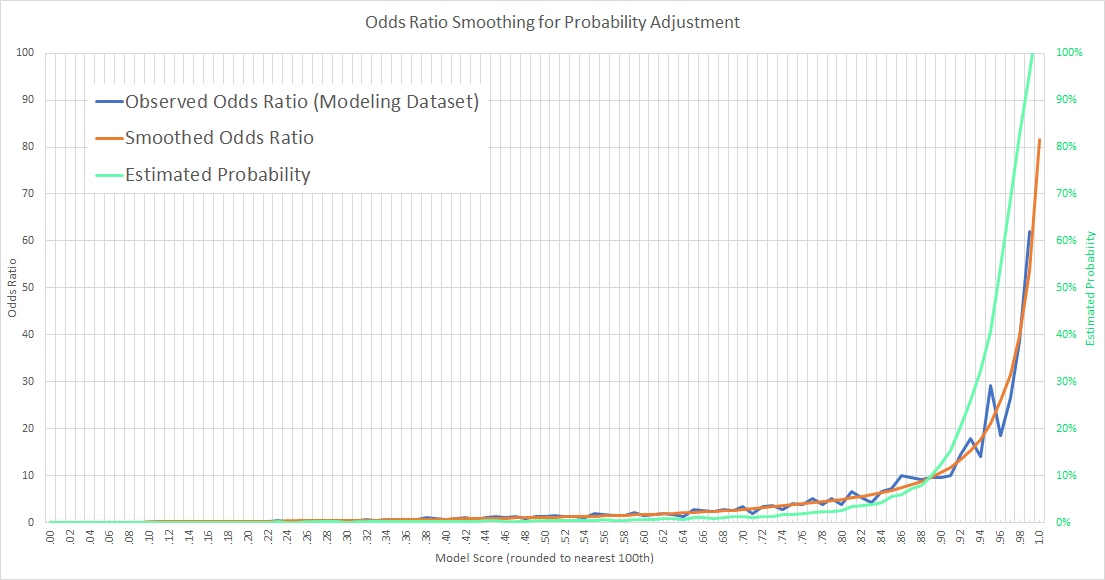

**Figure S6: Odds Ratio Smoothing.** The observed odds ratios from our modeling dataset was irregular, especially for model scores in the .90 to .98 range (blue line).  We engineered a simple smooth curve (orange line) for use in reversing the oversampled predicted probabilities from our regression model (green line).

**2. Implementing the model as a web-based query tool, and evaluating the performance of the model in that context.**

As mentioned in the Results section of the main text, the machine learning performance for the fitted model among the hold-out test cases, assuming a binary decision threshold of 0.5, was precision = 90.43% and recall = 84.57%, with F1 = 87.41%.  Overall accuracy was 87.81% with an AUC of 0.95. These results, particularly the AUC value, indicate that the model is inherently able to discriminate positive vs. negative examples quite well.

However, it is more relevant to evaluate the model in the context of the web-based ranking tool, which as implemented first identifies a pool of 5,000 candidate articles and then uses the model to make a ranked list according to their similarity scores.

**2.1 Creating a candidate pool of PubMed articles for matching to a given registered trial**

We devised a pre-selection methodology to reduce the number of candidate PubMed articles to those most likely to be associated with a specific trial.  First, we restricted the universe of possibly relevant PubMed articles to those containing the word “Trial” or “Trials” or “Multicenter Study” in title, abstract, or any metadata field, including publication type tags.  This yielded a subset of ~1.9 million articles.  Among this subset, we computed the number of trial condition words found within article titles, abstracts and MeSH terms as follows:

if Trial Conditions are MeSH terms, then add MeSH entry term synonyms, search for phrases, no stoplisting

if Trial Conditions are not MeSH terms, then conduct word search, employing the PubMed 365 word stoplist

if No Conditions are listed, then ignore and go on to Interventions

For trial interventions, we take all unique words found in the intervention XML fields from ClinicalTrials.gov and count the number of matching words in articles, excepting the PubMed 365 word stoplist.  We also calculated the total number of matching words from the trial title and description.

For a given registered trial, its candidate pool of PubMed articles are those that satisfy at least 1 condition match AND one intervention match.  To that list, we add several “catch-all” criteria derived from inspecting articles linked to trials that were missed due to the article title and abstracts never mentioning the exact condition and intervention as described in the trial record.  For example, articles related to hypertension, blood pressure, obesity, and “overweight” sometimes use popular phrasing not specifically considered to be MeSH terms, or synonyms.  Thus, we also include articles for which condition matches are greater than 2 OR intervention matches are greater than 3 or more than 32% of trial title and description words are found in article title, abstract, or MeSH.  If the resulting candidate pool contains > 5,000 articles, we sort the list by descending arithmetic average of (% conditions match, % interventions matched, % words matched) and keep the top 5,000.  We estimate that this preselection method captures 89.71% of the articles known to be linked to the trial (i.e., having NCT numbers) and 93.12% of such articles that are indexed as randomized controlled trials. Thus, the candidate pool provides very high recall while minimizing the search space severely (from 1.9 million to 5,000).  As described below, our online web tool also provides an “advanced search” mechanism that allows the user to bypass this pre-selection methodology to apply our model to an arbitrary PubMed query.

**3. ISSUES RELATED TO IMPLEMENTATION OF THE WEB-BASED TOOL**

**3.1 Using the Advanced query interface**

As mentioned in the main text, a publicly accessible web interface to this model may be found at http://arrowsmith.psych.uic.edu/cgi-bin/arrowsmith_uic/TrialPubLinking/trial_pub_link_start.cgi.  The default search strategy, i.e., the “Basic Search” setting, will launch the model application process for a preselected candidate pool of 5,000 articles.  Alternatively, the “Advanced Search” setting will present the user with PubMed query interface (Figure S7) which is pre-populated with a suggested PubMed query constructed from the publication types, trial start date, conditions, interventions, investigators, and other grant or project id’s from the indicated trial.  This query may be adjusted manually, or refined with PubMed.gov by clicking “View Results in PubMed.gov”.  Alternatively, the user may delete the entire query text and replace it with any valid query string for PubMed.gov, or a literal list of article PMIDs separated by spaces or commas.  Our website currently limits article sets to 100,000 articles, which are then processed as candidate articles and displayed as a ranked list in the same manner as in the Basic Search.

**Figure S7:  Our Advanced Search setting allows the site visitor to modify or paste-in any valid PubMed.gov article query or simply a list of article PMIDs, separated by spaces or commas.**

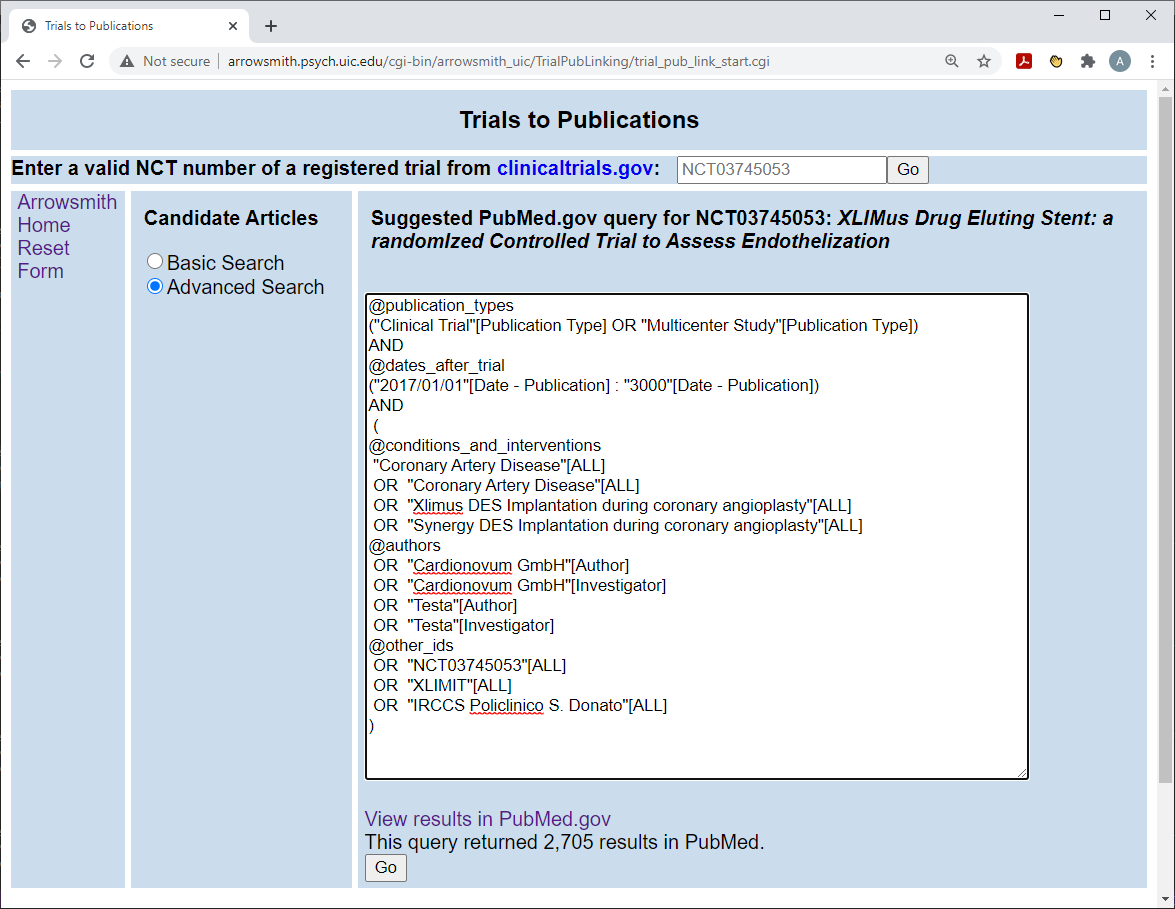

**3.2 International Registry Codes**

For any registered trial chosen by the user, our web tool displays a ranked list of articles. If an article is linked to any of a large number of International trial registries (see Table S8) we display all of its registry numbers. To do this, we developed python-flavor regular expressions for scraping international registry numbers from article metadata for display in our results page.  The Netherlands registry numbers were excluded from display because that registry’s previous use of the “NL” characters makes them indistinguishable from several substance names and genetic sequence monikers.  All of the international registry numbers other than clinicaltrials.gov are provided for information only and do not affect model calculation.

**Table S8: International registry codes scraped from article titles, abstracts and accession numbers.**

| Organization | Website | Registry Code Example | RegEx Search |
| --- | --- | --- | --- |
| World Health Organization – International Clinical | http://apps.who.int | U1111-1146-4747 | (U[0-9]+-[0-9]+-[0-9]+) |
| **ClinicalTrials.gov** | **https://www.clinicaltrials.gov** | **NCT02977780** | **(NCT[0-9]+)  case insensitive** |
| EU Clinical Trials Register | https://www.clinicaltrialsregister.eu/ | EudraCT2017-004065-27 | ([12][0-9]{3}-[0-9]{6}-[0-9]{2}) |
| Swiss National Clinical Trials Portal | http://www.kofam.ch | SNCTP000002431 | (SNCTP[0-9]+) |
| ISRCTN | http://www.isrctn.com/ | ISRCTN16230269 | (ISRCTN\s?[0-9]+) |
| Australian New Zealand Clinical Trials Registry | http://www.anzctr.org.au/ | ACTRN12619000738123 | (ACTRN[0-9]+) |
| Chinese Clinical Trial Registry | http://www.chictr.org.cn | ChiCTR1900023462 | (ChiCTR[0-9]+) |
| Clinical Trials Registry – India | http://ctri.nic.in/ | CTRI/2007/091/000017 | (CTRI\/[0-9]+\/[0-9]+\/[0-9]+) |
| Iranian Registry of Clinical Trials | http://www.irct.ir/ | IRCT138711051556N1 | (IRCT[0-9A-Z]+) |
| Clinical Research Information Service | https://cris.nih.go.kr | KCT0004066 | (KCT[0-9]+) |
| Philippine Health Research Registry | http://registry.healthresearch.ph/ | PHRR130117-000043 | (PHRR[0-9]+-[0-9]+) |
| Sri Lanka Clinical Trials Registry | http://www.slctr.lk/ | SLCTR/2019/022 | (SLCTR\/[0-9]+\/[0-9]+) |
| Thai Clinical Trials Registry | http://www.clinicaltrials.in.th/ | TCTR20190702004 | (TCTR[0-9]+) |
| Public Cuban Registry of Clinical Trials | http://registroclinico.sld.cu | RPCEC00000290 | (RPCEC[0-9]+) |
| Pan African Clinical Trials Registry | http://www.pactr.org/ | PACTR201808503792054 | (PACTR[0-9]+) |
| Tanzania Clinical Trial Registry | http://www.tzctr.or.tz/ | TFDA1900012 | (TFDA[0-9]+) |

Only NCT codes were used to determine gold standard trial-article pairs.  Additional information about international registries may be found at [www.hhs.gov](http://www.hhs.gov)
